# Healthcare worker intentions to receive a COVID-19 vaccine and reasons for hesitancy: A survey of 16,158 health system employees on the eve of vaccine distribution

**DOI:** 10.1101/2020.12.19.20248555

**Authors:** Michelle N. Meyer, Tamara Gjorgjieva, Daniel Rosica

## Abstract

Healthcare workers (HCWs) have been recommended to receive first priority for limited COVID-19 vaccines. They have also been identified as potential ambassadors of COVID-19 vaccine acceptance, helping to ensure that sufficient members of a hesitant public accept COVID-19 vaccines to achieve population immunity. Yet HCWs themselves have shown vaccine hesitancy in other contexts and the few prior surveys of U.S. HCW intentions to receive a COVID-19 vaccine report acceptance rates of only 28% to 34%. However, it is unknown whether HCW acceptance remains low following mid-November announcements of the efficacy of the first COVID-19 vaccines and the issuance of two emergency use authorizations (EUA) in December. We report the results of a December 2020 survey (*N* = 16,158; response rate 61%) administered by a large Pennsylvania health system to determine the intentions of its employees to receive a vaccine when it is offered to them. In a mixed sample of individuals serving in patient-facing and other roles, 55% would decide to receive a COVID-19 vaccine when offered, 16.4% would not, and 28.5% reported being undecided. The distribution of responses varied little across hospital campuses, between those in patient-facing roles and other HCWs, or by area or department of work. The higher rate of COVID-19 vaccine acceptance we observe may reflect the framing and timing of our survey. Among hesitant respondents, an overwhelming majority (90.3%) reported concerns about unknown risks and insufficient data. Other commonly reported concerns included known side effects (57.4%) and wanting to wait until they see how it goes with others (44.4%). We observed a substantial increase in self-reported intent to receive a COVID-19 vaccine after an FDA advisory committee voted to recommend an EUA. Among respondents who completed the survey after that point in time, 79% intend to receive a COVID-19 vaccine (*n* = 1155). Although only suggestive, this trend offers hope that rates of COVID-19 vaccine acceptance may be higher among HCWs and, perhaps, the general public than more hypothetical survey results have indicated.

## Introduction

Vaccines against COVID-19, the disease caused by the SARS-CoV-2 virus, present a critical milestone in the global response to the pandemic. But, as many have noted, vaccines don’t prevent disease; vaccinations do. Given the existence of vaccine hesitancy in other contexts (MacDonald et. al., 2015; Dubé et. al., 2013) and the fact that COVID-19 vaccines are, by definition, novel, there has long been concern by public health researchers and policymakers about whether sufficient numbers of people will be willing to take a COVID-19 vaccine to reach population, or so-called “herd,” immunity—estimated, in this context, to require vaccination rates of between 67% and 85% (Randolh & Barreiro 2020; Fontanet et al., 2020; Kwok et al., 2020; Higgins-Dunn & Lovelace 2020).

Several surveys of intention to receive a COVID-19 vaccine, when one becomes available, have been administered throughout the pandemic by academics, polling organizations, and market researchers. A metaanalysis of 30 such surveys resulted in an estimated global acceptance rate of 68.4% (Wang, W. et. al., 2020). Surveys of the U.S. population that asked respondents whether they would, would not, or were unsure about receiving a COVID-19 vaccine have reported rates of intention to receive a COVID-19 vaccine that range from 47% to 65%. Intentions have been somewhat volatile, perhaps reflecting the politicization of the pandemic response and election-related events.

The Centers for Disease Control and Prevention (CDC) has recommended that healthcare workers (HCWs)—”paid and unpaid persons serving in health care settings who have the potential for direct or indirect exposure to patients or infectious materials”—should be first to be offered a vaccine (along with residents of long-term care facilities), due to their increased exposure and their role in supporting critical healthcare functions (Dooling et al., 2020). Some survey evidence suggests that COVID-19 vaccine acceptance is greater among the general population if healthcare providers recommend it (Reiter et al., 2020). However, vaccine hesitancy in other contexts has been widely reported among HCWs (Paterson et. al., 2016).

Relatively few surveys of COVID-19 vaccine intentions have been conducted of HCWs, especially in the U.S. Surveys administered earlier in the pandemic, when vaccine enthusiasm was generally higher across the board (Gallup Panel, 2020; Morning Consult, 2020), found that outside the U.S., there was greater COVID-19 vaccine acceptance among HCWs than among the general population (Dror et al., 2020; Gagneux-Brunon et al., 2020; Harapan et al., 2020). More recent non-U.S. studies have found vaccine hesitancy among HCWs to be mostly in line with the general population (Grech et al., 2020; Wang, K. et al., 2020), jeopardizing hopes of clinicians serving as ambassadors of vaccine acceptance for their patients (Rowland, 2020).

In the U.S., the few surveys of HCWs that have been conducted have been administered later in the pandemic—but prior to public knowledge about specific COVID-19 vaccines—and have found that HCWs show equal or greater hesitancy than the general population. Two surveys conducted in October and October-November, respectively, found that only 34% of U.S. nurses (*N* = 12,939) would choose to be vaccinated (American Nurses Foundation, 2020) and only 31% of physicians and nurses (*N* = 370) would like to get the vaccine in the first wave (ReviveHealth, 2020), which is markedly lower than surveys of the general U.S. public conducted around the same time (Gallup Panel, 2020; Ipsos, 2020). A September survey found that just 28% of emergency medical services (EMS) personnel (*N* = 528) would choose to get the vaccine when it was available (Hatt, 2020) and a September-October survey found that only 32% of UCLA Health System employees (*N* = 609) intended to receive a vaccine as soon as possible (Gadoth et al., 2020)—again, notably lower than vaccine acceptance rates among the general public around the same time period (Tyson et al., 2020; Harris Poll, 2020: Hamel et al., 2020).

Reviewing the survey data published through December 10, a work group of the CDC Advisory Committee on Immunization Practices (ACIP) convened to consider the Pfizer-BioNTech COVID-19 vaccine concluded that overall acceptability of a COVID-19 vaccine was “moderate” (Oliver, 2020, slide 27). The ACIP noted, however, that most surveys were conducted prior to availability of specific information about the Pfizer-BioNTech COVID-19 vaccine. The companies announced on November 9 that an interim analysis suggested that their vaccine had an efficacy rate “above 90%” (Pfizer Inc., BioNTech, 2020). On November 18, they announced that their Phase III trial had met all of its primary efficacy endpoints and that an updated analysis found a 95% efficacy rate (Business Wire, 2020). To our knowledge, no published surveys of HCWs have been conducted since that date, and no surveys at all have been published that were conducted after the FDA issued an emergency use authorization (EUA) for the Pfizer-BioNTech vaccine on December 10. Yet intentions to receive a COVID-19 vaccine may be sensitive to vaccine-specific information such as efficacy and side effect profiles, to endorsements by local or national bodies, to the immediacy of the decision, and to social norms such as internal or external colleagues receiving the vaccine (or declining it).

Geisinger is a large, integrated health system serving central and northeast Pennsylvania. Beginning on December 4, as HCW decisions to receive or decline a COVID-19 vaccine appeared imminent, and to help inform the system’s plans for allocating and distributing limited supplies of COVID-19 vaccines across its 8 campuses, all employees were asked to complete a “COVID vaccine readiness survey” indicating whether they plan to receive a vaccine when one is available for them. The survey also sought to understand reasons for hesitancy, in order to inform internal communications and education efforts.

## Materials and methods

### Participants

A link to a short, 5-question survey hosted by Microsoft Forms was emailed to 26,361 Geisinger employees on December 4, 2020. Employees were told that the Pennsylvania Department of Health had directed that first doses of anticipated vaccines would be given to HCWs working in settings where symptomatic COVID-19 patients present for care, with other employees being offered a vaccine in phases over time. See the Discussion for a detailed description of the survey solicitation. The survey asked employees to identify the hospital campus closest to them; whether their role is clinical/direct patient-facing, not clinical/direct patient-facing, or other; and, for those with patient-facing or “other” roles, what area they work in (e.g., ICU, outpatient clinic, food services). Employees were then asked whether they would decide to receive the COVID-19 vaccine when one is available to them; those who responded “no” or “undecided” were then asked to select as many of the nine given reasons for their hesitancy that applied and/or to write in one or more additional reasons. Employees were asked to complete the survey by December 10, but the survey remained open and employees continued to submit responses. Reminders to take the survey were included in multiple issues of an internal daily newsletter distributed to all employees by email. Although this survey was designed and conducted as an administrative survey to inform operations and not as human subjects research, Geisinger’s IRB has determined that anonymous research surveys conducted by the authors are exempt from IRB review (IRB # 00008345).

### Free response coding

To analyze free responses, we used a content analysis approach and developed a ten-code codebook based on an initial review of a subset of the free responses. A coder then applied the codebook to categorize 1224 free responses provided by participants into the ten new categories. Free responses that simply recapitulated one of the nine pre-provided reasons were coded using the relevant pre-provided reason.

### Statistical analysis

All statistical analysis and data visualization was performed using R statistical software version 4.0.2 (R Project for Statistical Computing) and RStudio version 1.2.5001. Following evolving standards for reporting exploratory, non-confirmatory results (Benjamin et. al., 2018), we set statistical significance at α = 0.005 for 2-tailed tests; we describe results meeting a threshold of 0.05 as suggestive.

## Results

### Sample characteristics

A total of 16,158 Geisinger employees (61.3% response rate) have completed the survey at the time of writing. In order to keep the survey brief and to encourage honest responses, responses were anonymous and sociodemographic data were not collected. The average age of a Geisinger employee is 43 and employees span generational eras: 1% Silent Generation, 28% Baby Boomers, 22% Generation X, 46% Generation Y, and 3% Generation Z. Employees have worked for Geisinger for an average of 9.4 years. Employees are 73% female and 89% white.

The study sample included respondents who reported proximity to each of Geisinger’s eight campuses, with most respondents (44.1%) reporting closest proximity to Geisinger Medical Center (GMC), where the health system is headquartered (Figure 1A). Respondents work in a number of different areas. Of the 58% of respondents who work in a clinical or other direct patient-facing role and an additional 4.4% of respondents who did not describe their role as either patient- or non-patient-facing (Figure 1B), 28% work in ambulatory settings and clinics, 25.7% in inpatient care and another 6.6% in other inpatient settings, 7.3% in an ICU, and 6.1% in an emergency department (ED) (Figure 1C).

**Figure 1.**
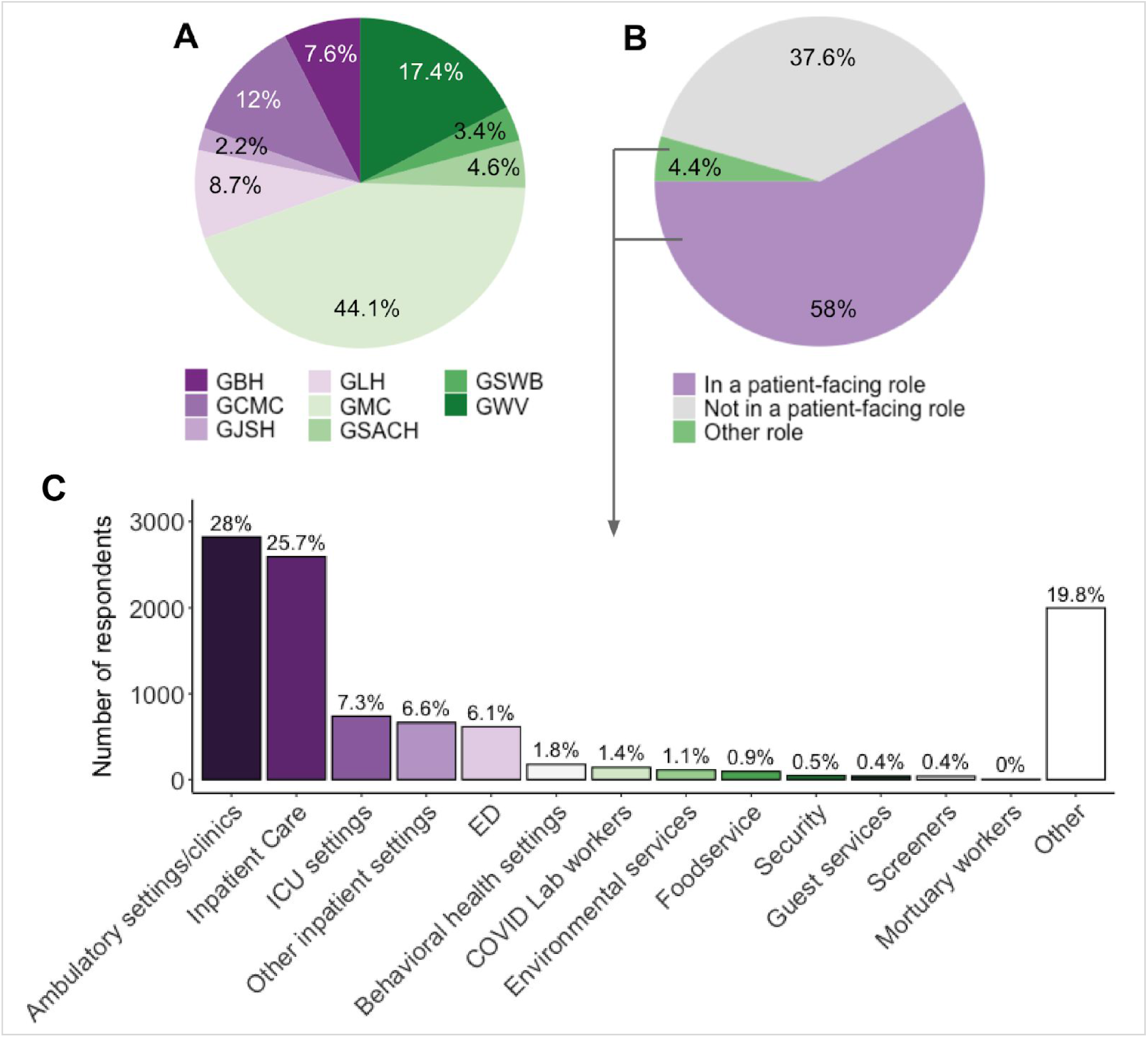
Sample characteristics. (A) Responses to question: “Which hospital campus is closest to you?”. (B) Responses to question: “Is your role clinical/direct patient-facing?. (C) Responses to question: “What area do you work in?” (shown only to respondents that have responded “Yes” or “Other” to the question in B).

### Respondents’ intent to receive a COVID-19 vaccine when available to them, by campus proximity, job role, and work area

When asked: “Will you decide to receive the COVID-19 vaccine when one is available to you?,” 55% of respondents said “yes,” 16.4% said “no,” and 28.5% were “undecided” (Figure 4A).

The distribution of responses varied little by proximity to different campuses: the percentage of vaccine-accepting respondents ranged only from a low of 51.5% nearest one campus to a high of 56.5% nearest another; the percentage of respondents who intend to decline a vaccine ranged from 14.9% to 19.6%; and the percentage of undecided respondents ranged from 27.5% to 33.4%.

Consistent with their increased exposure to the virus, respondents in patient-facing roles were more likely to say they intend to receive a vaccine (56.8%) than were those in non-patient-facing roles (51.4%, p<<0.005). However patient-facing workers were also slightly more likely to say they will decline a vaccine when one is available to them (17.5% vs. 15.6%, p<0.005). (Table S1). This may reflect the fact that patient-facing employees will also be offered a vaccine sooner, when less is known about the medium-term risks.

Among those in patient-facing roles, there were no clear or consistent patterns of intention across areas of work. Of those work areas with more than 100 respondents, the highest intent was reported among those working in ICU settings (63.1% of 738 respondents), inpatient care (60.9% of 2592), and other inpatient settings (60.5% of 661). Employees in these areas face greater exposure to the virus and, as a result, are among those employees who are the very first to be offered a COVID-19 vaccine. Yet the lowest intent was also reported among those working in emergency departments (51.9% of 616 respondents), who are also considered high-exposure and are in the same priority tier (Table S2). A limitation of these analyses is that a substantial number of respondents who were asked this question (19.8%) reported serving in an “other” work area (Figure 1C).

### Reasons for COVID-19 vaccine hesitancy

A total of 7265 respondents indicated either that they would decline a COVID-19 vaccine when one is available to them or that they were undecided (hereinafter, “vaccine hesitant” respondents). Vaccine hesitant respondents on average selected 2.73 ± 1.42 (mean ± s.d.) reasons for their hesitancy, with most selecting 3 reasons. An overwhelming majority (90.3%) of these respondents reported concerns about unknown risks of the vaccine, and more than half (57.4%) cited concerns about known side-effects such as headache and fatigue. A notable 44.4% reported that they would like to wait to take the vaccine until they see how it goes with others (Figure 2).

**Figure 2.**
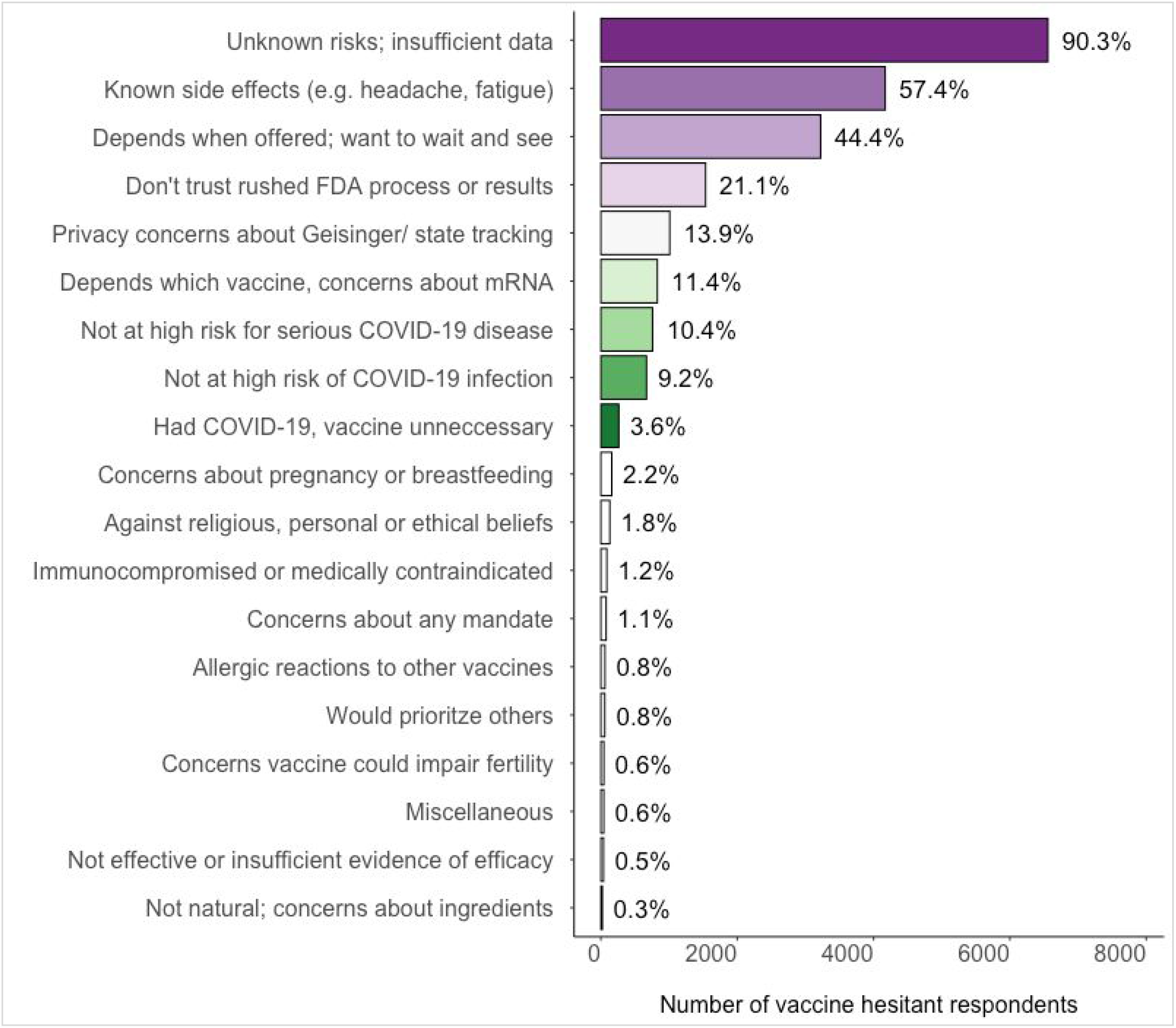
Respondents’ reasons for COVID-19 vaccine hesitancy

A total of 1224 respondents (16.1% of all hesitant respondents) provided additional reasons for their hesitancy beyond those provided in the survey instrument. These free responses were categorized based on a developed codebook (Methods) into ten additional codes. The most commonly raised concerns were those related to current or future pregnancy or breastfeeding (2.2% of the 7265 hesitant respondents). Other more frequently reported reasons included incompatibility with religious, personal or ethical beliefs (1.8%) and reportedly being immunocompromised or having another condition that made vaccination medically contraindicated (1.2%). A total of 77 respondents (about 90% of whom intend to decline a COVID-19 vaccine) spontaneously raised concerns about any potential COVID-19 vaccine mandate (1.1%). Other respondents reported having had allergic reactions to other vaccines (0.8%); raised concerns about the vaccine’s potential impact on fertility (0.6%); commented about the vaccine being ineffective or there being insufficient proof of efficacy (0.5%); indicated concerns that the vaccine’s ingredients were not natural (0.3%) or provided other miscellaneous reasons such as cost, fear of needles, or an aversion to being “experimented on’’ (0.6%) (Figure 2).

Compared to those who are undecided, those who intend to decline a vaccine were more likely to cite concern about known side effects (62% vs. 54.8%, p << 0.005), report distrust in the FDA process (36.3% vs. 12.2%, p << 0.005), and raise privacy concerns (22.9% vs. 8.7%, p << 0.005). Those who intend to decline a vaccine were also much more likely to select not being at high risk for either infection (17.5% vs. 4.4%, p << 0.005) or serious COVID-19 disease (20.5% vs. 4.6%, p << 0.005). Unsurprisingly, undecided respondents were much more likely to say that whether they will accept a vaccine depends on when one is offered to them, because they would like to “wait and see” how it goes with others (51.4% vs. 32.1%, p << 0.005). We also observed one suggestive difference between the groups, with those who intend to decline more likely to cite concerns about unknown risks or insufficient data than those who are undecided, p<0.05. (Figure 3A). Finally, respondents who intend to decline a vaccine were more likely to cite concerns related to pregnancy or breastfeeding (2.9% vs. 1.8%, p < 0.005); religious, personal or ethical beliefs (4.4% vs. 0.3%, p << 0.005); and concerns about a potential vaccine mandate (2.6% vs. 0.2%, p << 0.005).

**Figure 3.**
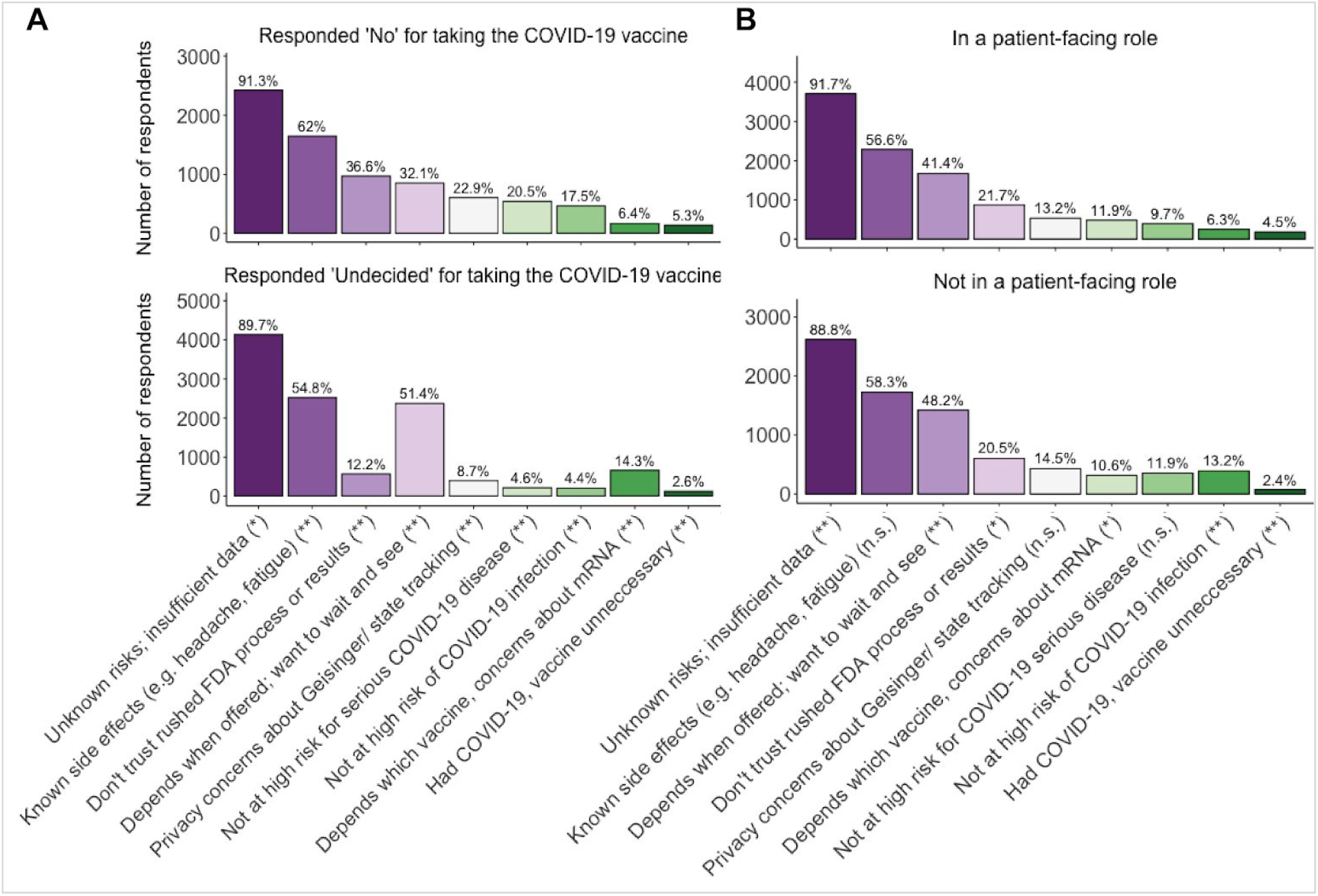
Reasons for COVID-19 vaccine hesitancy: comparison of those who intend to decline a vaccine versus those who are undecided (A), and of those in patient-facing roles versus those in non-patient-facing roles (B). p < 0.005 (**), p < 0.05 (*), p > 0.05 (n.s.)

Respondents serving in patient-facing roles were more likely to endorse concerns about unknown risks (91.7% vs 88.8%, p << 0.005) or reference having had COVID-19 as a reason for their hesitancy (4.5% vs. 2.4%, p << 0.005). Conversely, those in non-patient-facing roles were more likely to note that they would “wait and see” how it goes with others (48.2% vs. 41.4%, p < 0.005). We also found a few suggestive differences: those in patient-facing roles were more likely to distrust the FDA process (21.7% vs. 20.5%, p < 0.05) and more likely to report that their decision will depend on which vaccine they are offered (11.9% vs. 10.6%, p < 0.05) (Figure 3B).

We observe only a handful of significant or suggestive differences in the reasons for hesitancy across campuses. The largest difference regards concerns about known side effects: the frequency with which this reason for hesitancy was cited ranged from 50.8% among respondents who work closest to one campus to 67.4% among respondents who work closest to another campus, a difference in absolute percentage points of 16.6. Absolute differences in the frequency of other reasons by campus proximity ranged from 2.9% to 7.5%.

Somewhat more surprisingly, when we look at those areas in which at least 100 patient-facing respondents work, we similarly observe few significant or suggestive differences in the reasons for hesitancy (Figure S1). The largest difference is in concern about known side-effects: 48.9% of ICU workers endorsed this concern compared to 62.7% of those working in environmental services, with other work areas falling within that percentage point range of 13.8. Absolute differences in the frequency of other reasons by work area ranged from 3.1% to 10.8%.

### Intent to receive a COVID-19 vaccine, over time

During the period of data collection, a number of high-profile local, national, and international events related to the first administrations of a COVID-19 vaccine outside of clinical trials occurred that might plausibly have affected respondents’ intentions regarding COVID-19 vaccines. These events are described in the Discussion and some are plotted in Figure 4. To explore whether these events appear to have influenced HCW intentions to receive a COVID-19 vaccine, we first split the respondents into two subpopulations—those who submitted their survey response before the livestreamed VRBPAC vote to recommend an EUA on Dec 10, 2020, 5:38pm EST (*n* = 15,003), and those who submitted it at any time afterwards (*n* = 1128)—and compared the reported intentions between the two groups (hereafter, “pre-EUA” and “post-EUA” respondents). We found that, of the 7.2% of respondents who completed the survey post-EUA, 79% reported they would receive the COVID-19 vaccine; by contrast, of the 92.9% of respondents who had already completed the survey, 53.2% reported an intention to receive a vaccine. This difference was statistically significant (p << 0.005) (Figure 4B). We then looked at the distribution of responses per day and observed a steady increase in intent to receive a vaccine coinciding with several notable internal and external vaccine-related events (Figure 4C). Due to the considerable reduction in sample size over time and the possibility that latecomers to the survey are not representative, this finding is consistent with multiple interpretations and should be treated with caution.

**Figure 4.**
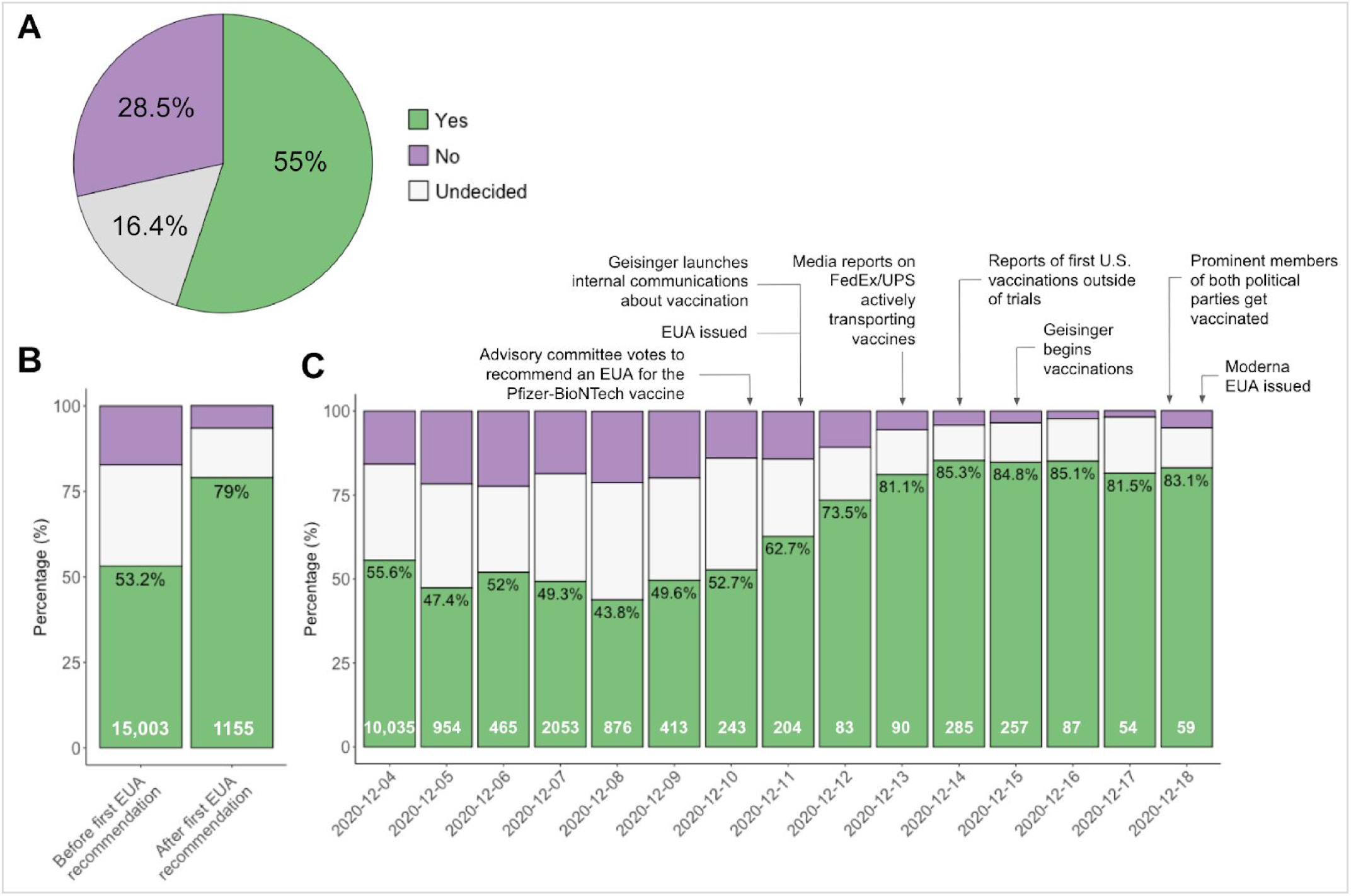
Intent to take the COVID-19 vaccine over time. (A) Responses to the question: “Will you decide to receive the COVID-19 vaccine when one is available to you?” (B) Intentions reported by those who submitted the survey before vs. after the VRBPAC vote to recommend the Pfizer-BioNTech vaccine EUA at 5:38 pm EST on December 10, 2020. (C) Intentions reported by those who submitted the survey on each of 15 days the survey has been open. Percentages in black refer to the percent of respondents reporting their intention to receive a vaccine. Numbers in white refer to the number of respondents who completed the survey during the relevant time period.

### Reasons for COVID-19 vaccine hesitancy, over time

Across all hesitant respondents, those who completed the survey pre-EUA were slightly more likely to say that whether they receive a vaccine will depend on when it is offered to them, because they would like to wait and see how it goes with others (57.3% vs. 53.9%, p < 0.05), as well as report feeling that they did not need a vaccine because they are not at high risk of infection (8.8% vs. 5.3%, p < 0.05). We observed no other significant or suggestive differences. Nor did we observe any significant or suggestive pre-vs. post-EUA differences in reasons for hesitancy among patient-facing or non-patient-facing respondents (or between them).

## Discussion

We surveyed over 16,000 employees of a large health system and found that 55% would decide to receive the COVID-19 vaccine when one is available to them. Although well below rates required for population immunity, this rate is substantially greater than those reported in similar surveys administered as recently as September to November, where reported intentions by HCWs to receive a COVID-19 vaccine ranged from just 28% to 34% (American Nurses Foundation, 2020; ReviveHealth, 2020; Hatt, 2020; Gadoth et al., 2020).

Several things might explain this observed increase in COVID-19 vaccine acceptance among the HCWs we surveyed. It could be that the population we sampled has always had substantially greater intentions to receive a COVID-19 vaccine than the other populations surveyed. For instance, surveys have consistently—and understandably—found greater COVID-19 vaccine hesitancy among members of the African American community (AP-NORC, 2020; Fisher et al., 2020; Gallup, 2020; Hamel, Lopes, Muñana et al., 2020; Kreps et al., 2020; Malik et al., 2020; Reiter et al., 2020; Tyson et al., 2020), and we surveyed a population that is 89% white. On the other hand, greater COVID-19 vaccine hesitancy has also been repeatedly reported among political conservatives and independents (Gallup, 2020; Hamel, Kearney, Kirzinger et al., 2020; Kreps et al., 2020; Morning Consult, 2020; Reiter et al., 2020; Tyson et al., 2020), and political conservatism is associated with residents of Geisinger’s catchment area in central and northeast Pennsylvania (WGAL, 2020).

It is also possible that the difference in rate of vaccine acceptance that we observe at least partially reflects the different posture of our survey. Almost all other surveys were administered by polling organizations or academic researchers and designed simply to measure national opinion. Our survey, by contrast, was administered by an employer tasked with distributing vaccines to interested respondents. The survey was administered via a relatively rare system-wide “administrative announcement” email and was attributed to Geisinger’s Division Chief of Infectious Diseases, who stated, “we encourage you to strongly consider taking the vaccine.” The source of information, such as an authority figure, has been shown to influence the extent to which recipients incorporate the information into their decision-making (Dolan et al., 2012).

Although our survey, like the others, measured only intentions and not behaviors, our survey framed the decision whether to receive a COVID-19 vaccine as imminent for respondents. Stated intentions and actual behaviors are known to frequently diverge (Sheeran & Webb, 2016), and our survey may have measured intentions in a way that helped close that gap to some extent. For instance, the survey was described as a “COVID-19 Vaccination Readiness Survey,” employees were told that Geisinger expected that multiple vaccines would “soon receive” an EUA, that Geisinger was “actively planning” for distribution among employees, “potentially in the next few weeks,” and that respondents’ “time-sensitive response [was] needed” to help effectively plan that distribution process.

The announcement used a gain frame, noting that the vaccines were expected to be as much as 90 to 95 percent effective, and further contextualized this information by noting that COVID-19 vaccines are therefore “substantially more effective than the flu vaccine” (which Geisinger employees are required to take). Gain-framed messages have been shown to be effective in promoting certain prevention behaviors (O’Keefe & Jensen, 2007). Immediately after noting the vaccines’ substantial benefits, employees were told that initial supplies would be “very limited.” When a resource is in short supply, desirability for that item tends to increase (Cialdini, 2001) and limited-quantity scarcity messages have been found to positively influence consumption intentions (Aggarwal et al., 2013).

It seems likely that the overall increased rate of COVID-19 vaccine acceptance we observe compared to Fall 2020 surveys is also at least partly due to the more recent timing of our survey. Between fall and early December, new material information about vaccines became available, such as the very high vaccine efficacy rates and reassuring short-term safety profiles of the Pfizer-BioNTech and Moderna vaccines; as discussed above, the efficacy information was explicitly incorporated into our survey solicitation message. Our respondents may also have been influenced to have a more favorable view of the benefits of COVID-19 vaccines in light of the recent national surge in COVID-19 cases, hospitalizations, and deaths (New York Times, 2020), which Geisinger has also experienced.

Even within the period of our December data collection, we observe a trend over time that suggests still-increasing COVID-19 vaccine acceptance. This could reflect several events that occurred during this time. Late on December 10, the Vaccines and Related Biological Products Advisory Committee (VRBPAC) to the FDA voted to recommend an emergency use authorization (EUA) for the Pfizer-BioNTech COVID-19 vaccine (Stat Staff, 2020). The next morning, the FDA announced that it would issue an EUA shortly (Hahn & Marks, Dec. 11, 2020), and it did so late that day (Hinton, 2020). At that point, Geisinger launched an internal informational campaign about the vaccine and its imminent distribution, some materials of which are public (Geisinger, 2020). Internal materials include video testimonials from employee leaders who explain why they have chosen to receive the vaccine. On December 12, the ACIP voted to recommend the vaccine (Oliver et al., 2020), and the next day, national media reported images of FedEx and UPS vehicles transporting the vaccine, accompanied by U.S. Marshals and, often, cheering onlookers. One video of trucks leaving Pfizer’s facility in Kalamazoo, Michigan, was posted to Twitter and has been viewed over 4 million times (Muntean, 2020). On December 14, media published the images of HCWs receiving the vaccine. Vaccination of HCWs began at Geisinger on December 15. Late on December 17, the VRBPAC voted to recommend an EUA for Moderna’s COVID-19 vaccine (Stat Staff, 2020), the FDA indicated that it would issue an EUA shortly (Hahn & Marks, Dec. 17, 2020). On December 18, several prominent members of both major political parties, including Vice President Mike Pence (whose vaccination was carried live on major news networks, including Fox News), Second Lady Karen Pence, Democrat House Speaker Nancy Pelosi, and Republican Senate Majority Leader Mitch McConnell—but, conspicuously, not President Trump—publicly and enthusiastically received the Pfizer-BioNTech vaccine (Kraft-Todd et al., 2020; Peters, 2020). That evening, the FDA issued an EUA for the Moderna vaccine and President-Elect Joe Biden issued a statement saying that he looked forward to receiving a vaccine on Monday, December 21, and set a goal for administering 100 million vaccine shots in the first 100 days of his administration (Biden, 2020). As the vaccination rollout got under way in earnest, some patient-facing HCWs at other health systems publicly protested their systems’ vaccine allocation schemes, which they felt unfairly deprioritized them relative to their exposure to the virus (Bernstein et al., 2020; Eisenberg, 2020).

Although caution is warranted in interpreting our observed day-by-day increase in vaccine acceptance as representative of our sample population, these events could have helped associate COVID-19 vaccines with several sentiments that should increase intention to receive a vaccination. These include: the sense the receiving a COVID-19 vaccine is normative among influential people and/or HCWs (Cialdini, 2007; Goldstein et al., 2008; Salmon et al., 2014; Venema et al., 2020); that, perhaps unlike some other pandemic behaviors, vaccination is bipartisan; that in receiving a COVID-19 vaccine, especially in the first wave, one participates in an historic moment; and that early vaccination is a reflection of the importance of one’s heroic role on the front lines of the pandemic response, such that being deprioritized for vaccination is, in the words of one healthcare worker, “insulting” (Bernstein et al., 2020).

We observe a significant, but surprisingly modest, increased intention to receive a vaccination among patient-facing HCWs and, within patient-facing employees, respondents who work in areas of the highest exposure are both most and least likely to say they will receive a vaccine.

### Limitations

Our survey has several limitations. Because the survey was anonymous, we cannot exclude the possibility that some respondents took the survey more than once. They might have done so if they believed that the survey results could affect availability of a vaccine at Geisinger, their nearest campus, or for themself. Respondents live and work in one particular region of the U.S. and results might not be generalizable to other U.S. HCWs. Our analyses of intention over time is limited by the fact that latecomers to the survey might not be representative of the sampled population as a whole. Finally, we asked respondents whether they intended to receive a COVID-19 vaccine when one is available to them, not ever, so rates of decline should be interpreted accordingly.

## Data Availability

Data, materials, and code are available upon request to the corresponding author.

## Acknowledgements

We thank the Geisinger employees who responded to the survey. We also thank Stephanie Gryboski, MHA, Allison Hess, MBA, and Stanley Martin, MD, for input into the survey instrument. We thank Geisinger Marketing for administering and promoting the survey. Finally, we thank participants in “Behavioral Science and COVID-19 Vaccine Acceptance,” a virtual meeting convened by the University of Pennsylvania on December 16, 2020, for helpful comments on a presentation of some of these results.

## Supplementary Information

**Table S1.** Intent to take COVID-19 vaccine, by type of role

| Intent to receive<br>COVID-19 vaccine | N | Percentage (%) |
| --- | --- | --- |
| **In a clinical/patient-facing role** |  |  |
| Yes | 5328 | 56.8 |
| No | 1637 | 17.5 |
| Undecided | 2408 | 25.7 |
| **Not in a clinical/patient-facing role** |  |  |
| Yes | 3117 | 51.4 |
| No | 948 | 15.6 |
| Undecided | 2005 | 33 |
| **Other role** |  |  |
| Yes | 448 | 62.7 |
| No | 71 | 9.9 |
| Undecided | 196 | 27.4 |

**Table S2.**
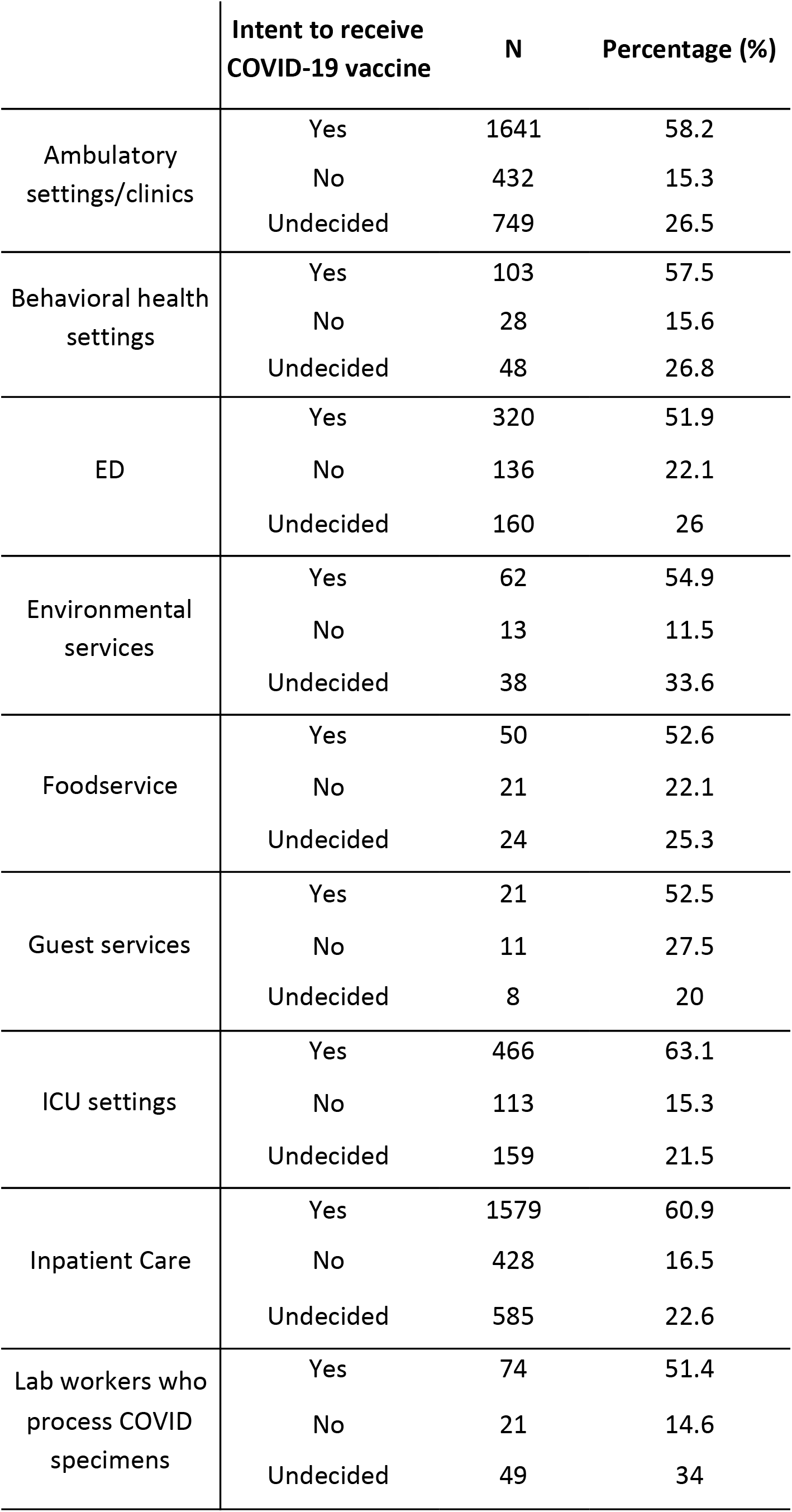
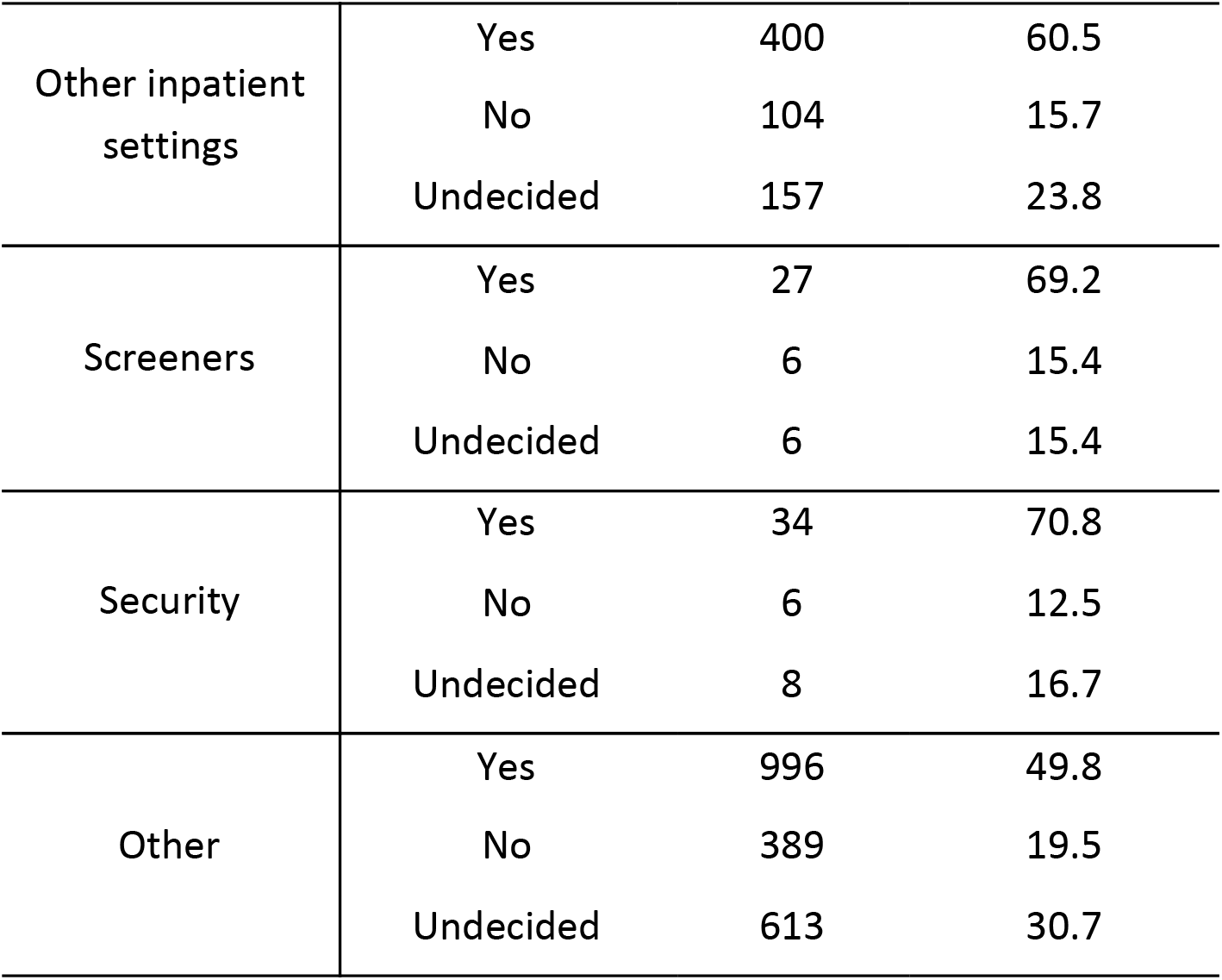
Intent to take COVID-19 vaccine, by area of work

**Figure S1.**
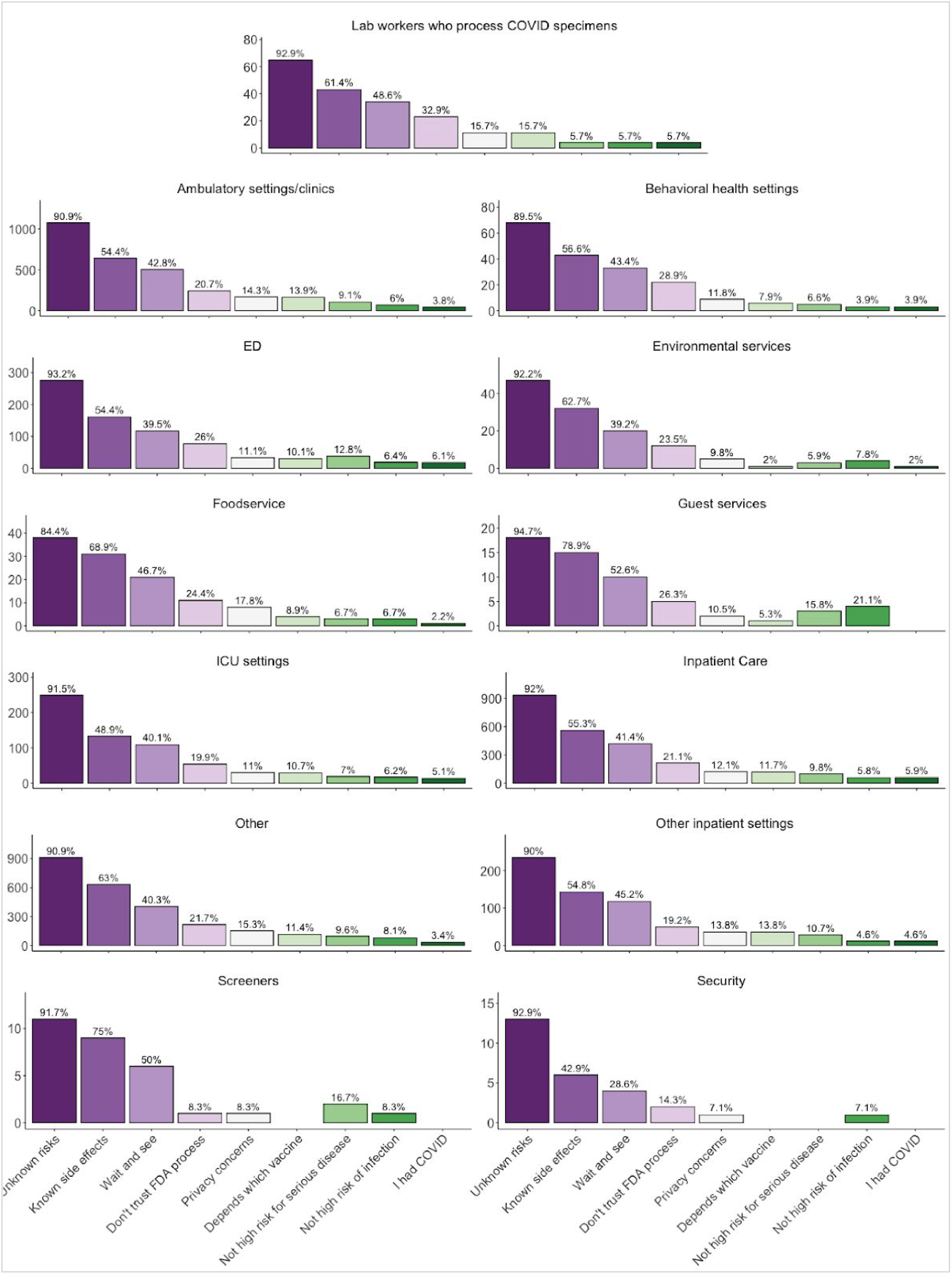
Reasons for COVID-19 vaccine hesitancy, by area of patient-facing work

